# Epidemiological Insights and Duffy Binding Protein Evolution of *Plasmodium vivax* in Duffy-Negative Cameroonians

**DOI:** 10.1101/2025.05.29.25328521

**Authors:** Cheikh Cambel Dieng, Rene Teh Ning, Canelle Kipayko, Regan Schroeder, Nontokozo Mdluli-Berndt, Bate Ayukenchengamba, Zidedine Nematchoua, Sona Doris, Ambendekson Elizabeth Reward, Irene Sumbele Ngole Ule, Helen Kuokuo Kimbi, Eugenia Lo

## Abstract

Malaria remains a major public health concern in sub-Saharan Africa, and *P. vivax* is emerging in regions with predominantly Duffy-negative populations. This study investigated *P. vivax* prevalence and genetic diversity across three ecological zones in Cameroon. *P. vivax* was detected in ∼11% of febrile patients and 5.5% of asymptomatic individuals, all are Duffy-negatives. Clinical infections exhibited higher parasitemia than infections detected in the communities. Standard PvLDH-based RDTs produced false-negative results, even at high parasitemia levels, whereas molecular tools like qPCR demonstrated superior sensitivity. Genetic analysis of *PvDBP1* identified two prevalent mutations, I379L (73%) and E225K (61%), among samples, suggesting adaptive evolution. Phylogenetic analysis clustered Cameroonian isolates with those from Botswana but distinct from the Ethiopian and Sudanese isolates, indicating historical migration and local adaptation. The detection of asymptomatic *P. vivax* cases highlights the potential for transmission, reinforcing the need for enhanced surveillance in both community and clinical settings.

## Introduction

Malaria remains a significant global health challenge in sub-Saharan Africa. For decades, *Plasmodium vivax* (*Pv)* was thought to be rare in African populations that lack the Duffy blood group antigen expression. However, emerging evidence has identified *Pv* cases in Duffy-negative individuals across Africa, challenging this conventional dogma. The endemic range of *Pv* has extended beyond the Horn of Africa and penetrated Duffy-negative regions (>95%). A point mutation (c.1–67T>C; rs2814778) in the GATA-1 transcription factor binding site of the Duffy antigen/receptor for chemokines (DARC) gene promoter alters erythroid expression, reducing Duffy antigen expression on the surface of the red blood cells (1, 2).

In sub-Saharan Africa, malaria prevalence varies significantly by region and season. For instance, in Cameroon, the highest prevalence occurs in the western and northern regions that are mostly highlands, which could exhibit different malaria transmission dynamics compared to the lowlands in the eastern region (3). Malaria prevalence also varies significantly by season, ranging from 30% in the dry season from November to February to 65% during the rainy season from March to October. The western part of Cameroon experiences heavy seasonal rainfall, which drives malaria transmission. While *P. falciparum* (*Pf*) remains the dominant parasite species, *Pv* cases have been reported among travelers who acquired infections in Cameroon and were diagnosed after returning to their home countries (4, 5). “*Pv*-like” asymptomatic infections have also been detected by microscopy in schoolchildren in the Mount Cameroon area. Two recent studies also reported an infection rate of 5.6% (6) and 35% (7) of *Pv* in febrile Duffy-negative individuals in Dschang, northwestern Cameroon by PCR. However, most of these studies were case reports and did not assess the broader epidemiological or genetic characteristics of *Pv* in the region. The extent of genetic polymorphisms of the Duffy Binding Protein (DBP) gene, a key gene that the parasites use to invade human erythrocytes via DARC (8), is unclear among *Pv* from Duffy-negative populations in Central Africa. Such variations may influence the adaptability and geographic spread of *Pv* across Africa.

Environmental factors such as temperature, rainfall, and elevation can shape *Pv* transmission dynamics through their effects on mosquito vectors and parasite development. Across Cameroon’s diverse ecological zones, these conditions may influence regional patterns of *Pv* transmission (9). In this study, we assessed *Pv* prevalence among the northwest, southwest, and eastern regions of Cameroon in both community and hospital settings. We further examined *Pv* genetic polymorphisms from these regions and compared them to isolates from East Africa and beyond, to evaluate genetic diversity and infer evolutionary relationships. Understanding the epidemiological and genetic features of *Pv*, particularly in Duffy-negative populations, is essential for informing malaria surveillance and control strategies in Africa.

## Methods

### Ethical Statement

The study protocol was approved by the Institutional Review Boards of the Universities of Buea and Bamenda (Ref No. 2024/2110-06/UB/SG/IRB/FHS). Informed consent was obtained from all participants or their legal guardians. All methods were carried out in accordance with relevant guidelines and regulations.

### Study Sites and Sample Collection

A cross-sectional study was conducted across three study sites in Cameroon including Bamenda, Buea, and Bertoua. Site selection was based on variation in climate and landscape. Bamenda lies in a highland region (1201–1400 m). Buea, at 530 meters elevation, is a multi-ethnic semi-urban area with year-round malaria transmission (10, 11). Bertoua, in the Eastern region, has a wet equatorial climate and is known for timber and mining industries (9). A total of 1,373 samples (408 samples from Buea, 498 from Bamenda, and 467 from Bertoua) were collected between April and August 2023, of which 793 were from febrile patients at health facilities and 580 from community households. Among all samples, 555 were males and 818 were females. For each individual, 30–50 μL of finger-pricked blood was spotted onto Whatman 3MM filter paper, air-dried, and stored until DNA extraction. Thick and thin blood smears were also prepared for microscopic screening of *Plasmodium*.

### *Plasmodium* Screening, Duffy Genotyping, and *PvDBP1* Sequencing

*Plasmodium* species were initially determined using rapid diagnostic tests (RDTs; Bioline Malaria Ag Pf/Pv manufactured by Abbott Inc) specific to *Pf* (PfHRP2) and *Pv* (PvLDH), as well as microscopy. Molecular assays were conducted for further diagnosis. Parasitic DNA was extracted from dried blood spots using a modified Chelex®-saponin method (12, 13).

Quantitative PCR (qPCR) targeting the 18S rRNA gene was used to identify mono-*Pv* and mixed *Pv-Pf* infections (14). Parasitemia was measured via SYBR Green detection method, using *Pv*-specific primers (S1Table) (13). Each run included positive controls from *Pv* Pakchong (MRA-342G) and Nicaragua (MRA-340G) isolates, along with negative controls. A standard curve was generated based on a ten-fold serial dilution of a *Pv* control plasmid to assess amplification efficiency (E). Specificity of the amplification was verified by melting curve analysis. For each sample, the mean cycle threshold (Ct) value and standard error were calculated from three independent runs. Parasitemia was determined using the equation: 2^E × (40 - Ct__sample_).

For each sample, *PvDBP1* region II was also amplified using published primers (forward: 5′-GATATTGATCATAAGAAAACGATCTCTAGT-3′; reverse: 5′-TGTCACAACTTCCTGAGTATTTTTTTTAGCCTC-3′) (15), designed based on the *P. vivax* reference sequence PVX_110810 from the Sal I strain (NC_009911.1). The amplification was performed following the published protocol (15). PCR products were purified prior to Sanger sequencing. For host Duffy genotyping, a TaqMan qPCR assay was used to examine the point mutation (c.1–67T>C; rs2814778) of the *DARC* gene (16) . A no-template control was used in each assay. Genotypes were determined by the allelic discrimination plot. In addition, a 1,100-bp fragment of the *DARC* gene was further amplified for a subset of samples using published primers for Sanger sequencing to confirm Duffy genotypes (17).

### Data analysis

To evaluate factors associated with mono-*Pv* and mixed *Pv-Pf* infections, odds ratios (OR) and 95% confidence intervals (CI) were calculated for various demographic, clinical, and epidemiological variables. Demographic variables including gender (male vs. female), age (<5 years, 5–15 years, and >15 years), and collection origin (community vs. hospital-based) were analyzed to examine the potential risk factors for infection. Symptom-based analyses were conducted to assess differences in clinical presentation across infection types. MedCalc software was used for all statistical analysis.

*PvDBP1* sequences were aligned to the P01 (PlasmoDB-67_PVP01) reference strain using MUSCLE in Geneious vX.1. Codon mutations in the Cameroonian *Pv* were compared with isolates from Thailand and China (Asia), Brazil (South America), Sudan and Ethiopia (East Africa), and Botswana (Southern Africa) that represent geographically and epidemiologically diverse regions. Phylogenetic trees were constructed using Maximum Likelihood (ML) method in RAxML. The ML tree was generated with 1,000 bootstrap replicates. The General Time Reversible (GTR) model with gamma distribution was used to account for rate heterogeneity.

The resulting tree was imported into FigTree v1.4.4 for visualization. Sequences are available under GenBank submission number 2920113.

## Results

### Prevalence of *Pv* infections among Duffy-negatives

The prevalence of mono-*Pv* and mixed *Pv-Pf* infections varied significantly across study sites (Figure 1). Among 793 hospital samples, mono-*Pv* prevalence was 11% (n=86), mono-*Pf* was 24.5% (n=192), and mixed *Pv-Pf* infections were 13.5% (n=106; Figure 2A, Table 1). All *Pv* infections were Duffy-negative (CC) per TaqMan genotyping. The highest rates of mono-*Pv* and mixed *Pv-Pf* infections were found in females over 15 years old in Bamenda (6% and 3.8%).

**Figure 1.**
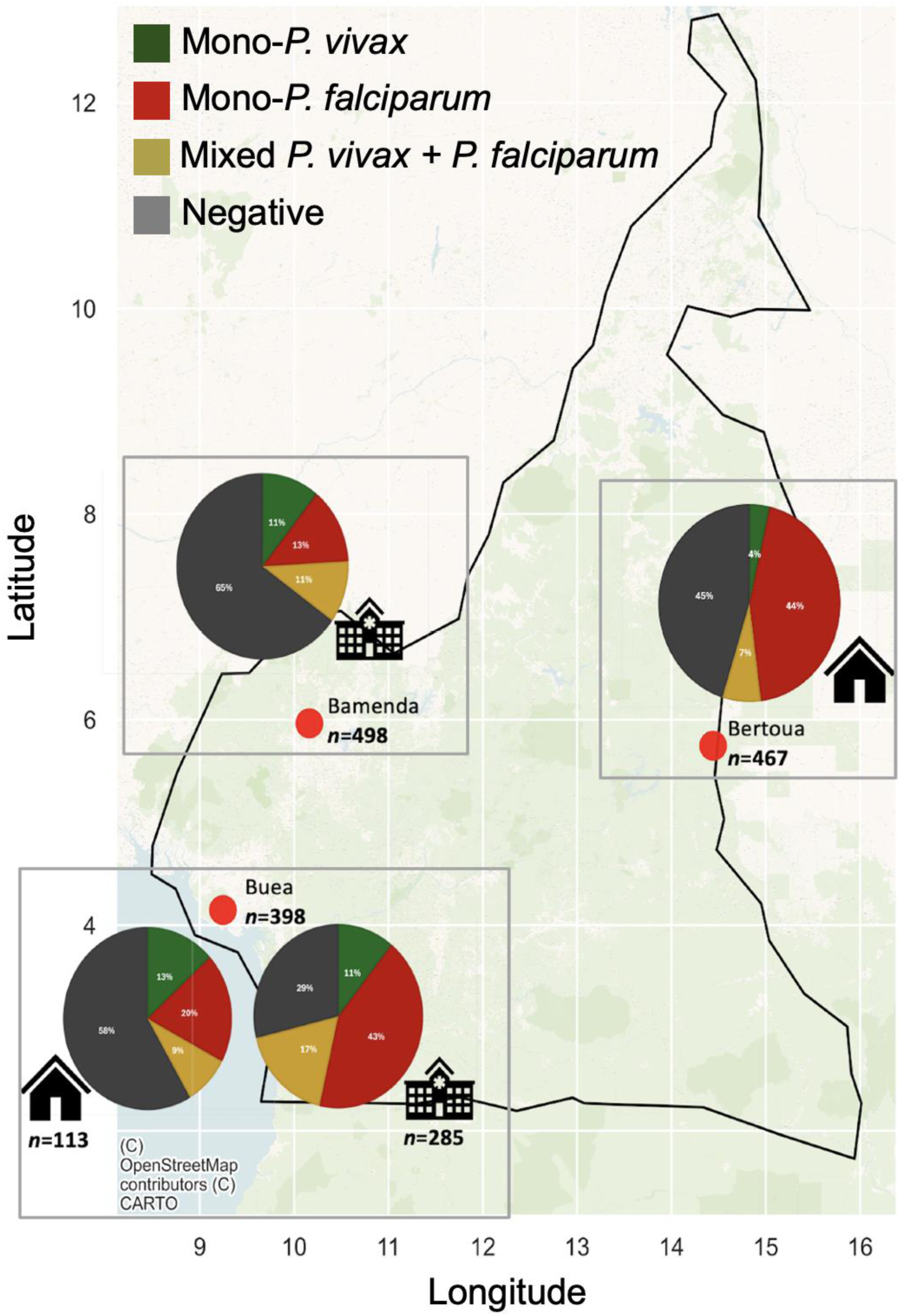
Study sites across Cameroon (Bamenda, Buea, and Bertoua) by hospital and community-based collection and infection type including mono-*P. vivax*, mono-*P. falciparum*, mixed *P. vivax* and *P. falciparum*, and negative samples. The prevalence of mono-*P. vivax* and mixed *P. vivax*-*P. falciparum* infections showed significant variation across the study sites.

**Figure 2.**
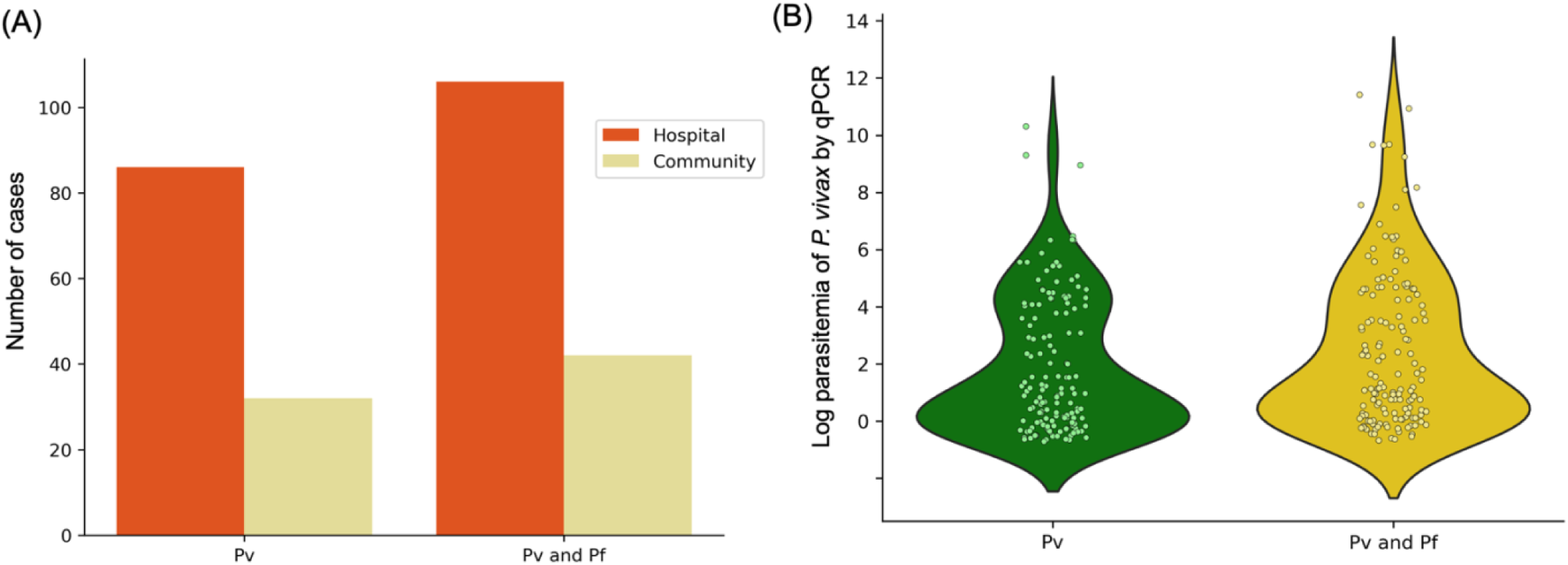
Comparison of *Plasmodium vivax (Pv)* and *Plasmodium falciparum (Pf)* Infections Across Hospital and Community Settings and Parasitemia Levels in Mono and Mixed Infections. (**A**) Panel shows the number of cases for *P. vivax* and mixed *P. vivax* and *P. falciparum* infections across hospital (orange bars) and community (olive green bars) settings. Among the hospital samples, 10% (86) were positive for *P. vivax* while 52.3% (409) samples showed mixed infections, with both *P. vivax* and *P. falciparum* present. In community samples, 5.5% (32) were positive for *P. vivax* and 47.8% (277) samples showed mixed infections.

**Table 1.**
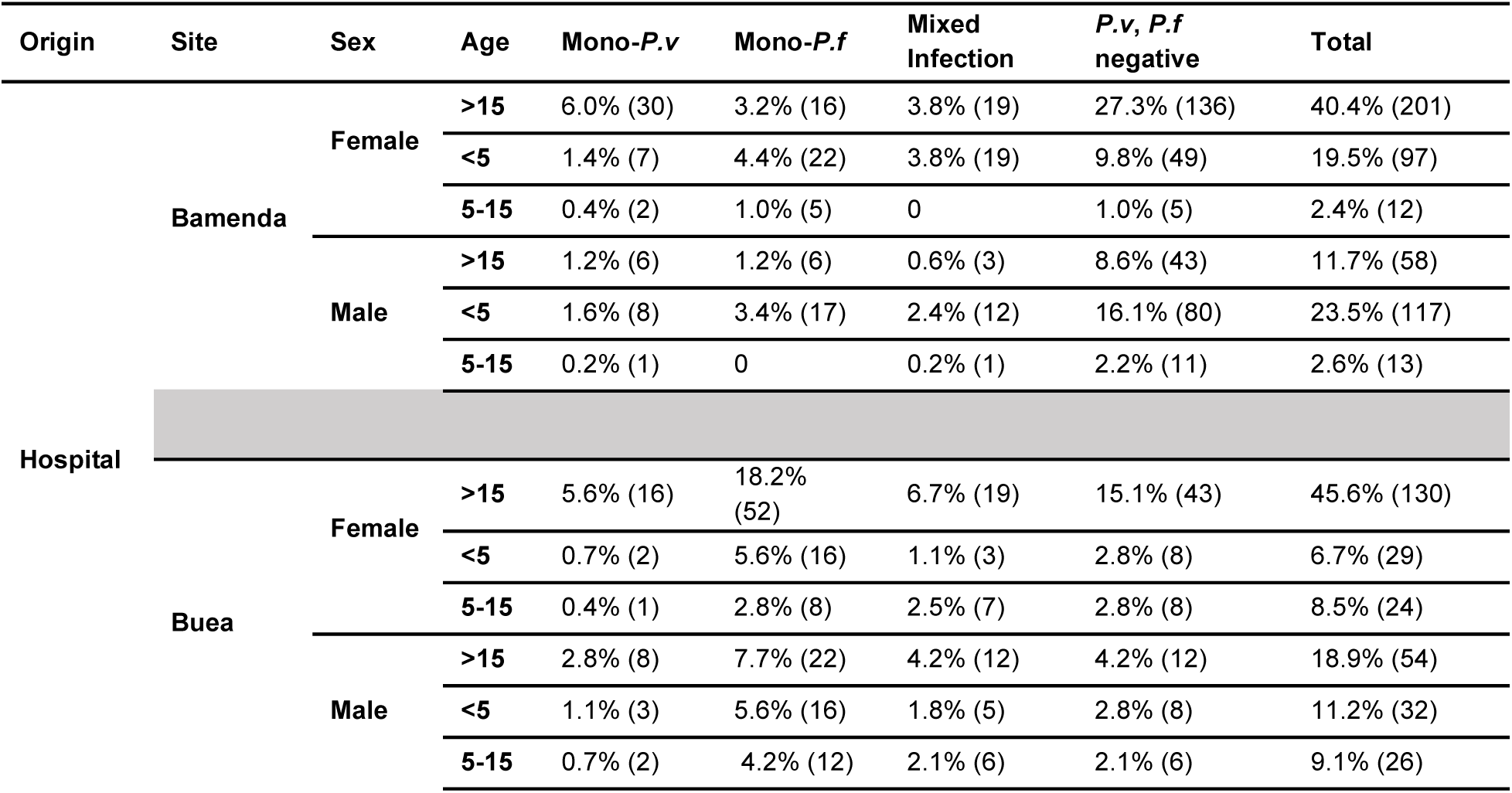
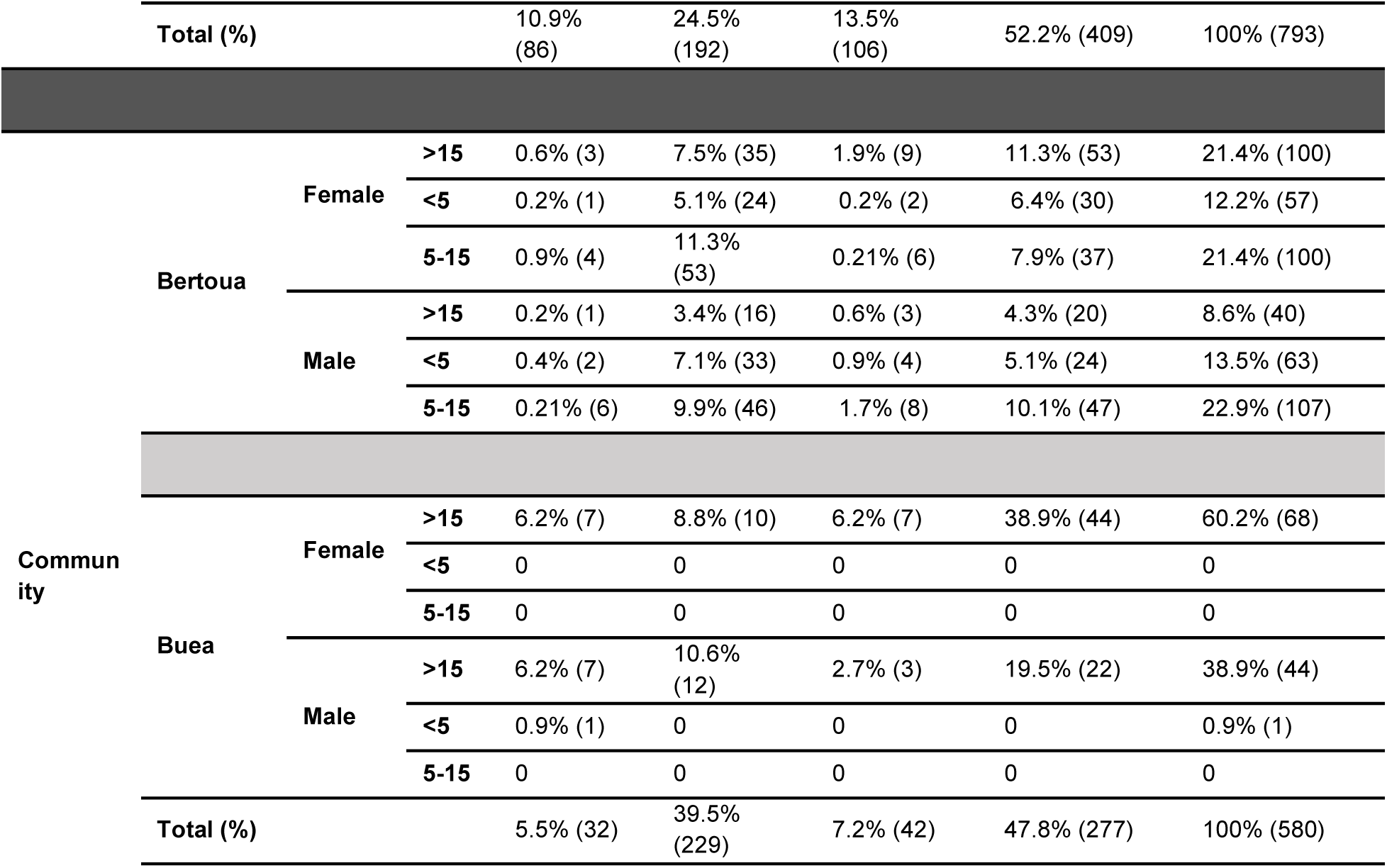
Prevalence of Mono-*Plasmodium vivax*, Mono-*Plasmodium falciparum*, and mixed infections across different sex and age groups in Bamenda, Bertoua and Buea. This table summarizes the infection prevalence of Mono-*Plasmodium vivax* (Mono-Pv), Mono-*Plasmodium falciparum* (Mono-Pf), and Mixed Infections across three sites in Cameroon (Bamenda, Bertoua, and Buea). Data are stratified by sex and age groups (>15 years, <5 years, 5-15 years). For each site, the total number of cases and the percentage distribution of infections are provided. Chi-square tests were conducted to assess the association between infection type (Mono-Pv, Mono-Pf, Mixed) and demographic variables (age group, sex). Significant differences were observed in infection prevalence among different age groups, with higher mixed infection rates observed in females, especially children under 5, indicating a greater vulnerability to mixed infections (p-value < 0.05). Specifically, the highest prevalence of mixed infections was observed in Bamenda (36.54%), followed by Bertoua (34.26%) and Buea (29.20%).

Among 580 community samples, mono-*Pv* prevalence was 5.5% (n=32), mono-*Pf* was 39.5% (n=229), and mixed *Pv-Pf* infections were 7.2% (n=42). *Pv* infection rates were comparable between females (8.9%) and males (8.1%, OR=1.11, *p*=0.597). Similarly, mixed infections occurred in 11.1% of females and 10.3% of males (OR=1.09, *p*=0.617). No significant differences in parasitemia were observed between mono- and mixed infections, sexes, or age groups (Figure 2B; S1 Fig.; S2 Fig.).

Of 119 *Pv*-positive samples by 18S-qPCR, 112 (94%) were confirmed by *PvDBP*1-based PCR. However, all qPCR-positive samples were microscopy-negative, and only 15 (12.6%) were RDT-positive (Table 2). These 15 RDT-positive samples showed relatively low Ct values (i.e., high parasite DNA copies) by qPCR (Table 2). These findings highlighted the limited sensitivity of conventional diagnostics in detecting *P. vivax* infections, particularly in Duffy-negative individuals. While clinical samples had significantly higher average parasitemia than community samples (*p*<0.001; Figure 3), several community-based (asymptomatic at the time of collection) infections were shown with relatively high parasitemia. Specifically, 13% of community samples had parasitemia >1,000 parasites/μL, compared to 21% of hospital cases showing such level of parasitemia.

**Figure 3.**
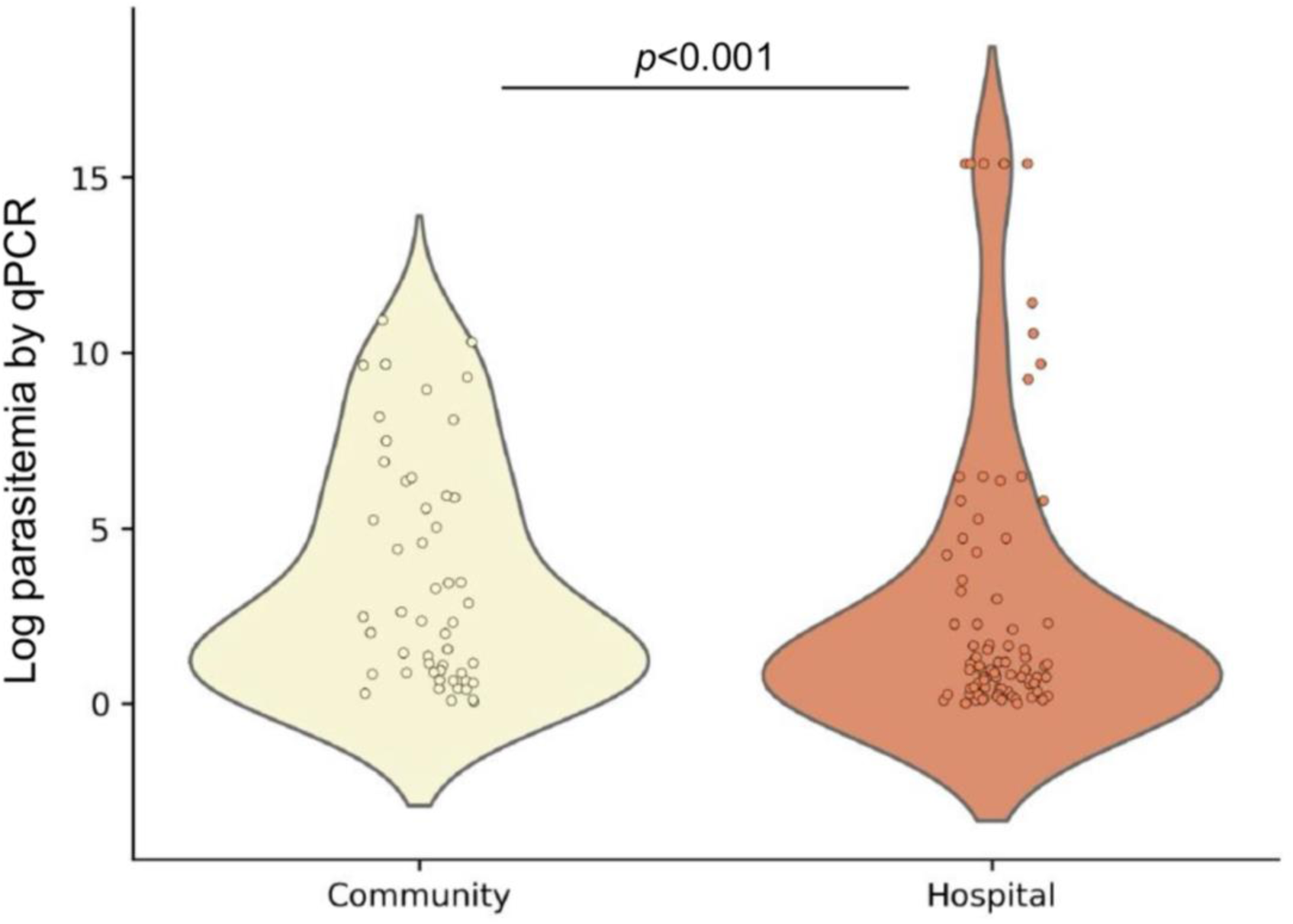
Comparative Analysis of Parasitemia Levels by Sample Origin in *P. vivax* Infections. Compares parasitemia levels between hospital and community samples, showing significantly higher parasitemia in hospital samples.

**Table 2.** Detection of *Plasmodium vivax* Infections across Hospital- and Community-Based Samples. This table shows the *P. vivax* detection using 18S-qPCR, *PvDBP1*-targeted PCR (DBP-PCR), microscopy, and rapid diagnostic tests (RDTs) in hospital-based (N = 793) and community-based (N = 580) samples. Most infections, particularly those with higher Ct values (indicative of lower parasite densities), were undetectable by microscopy and RDT. All qPCR-positive *P. vivax* samples were from Duffy-negative individuals.

| Hospital-based samples (total N= 793) |  |  |  |  |
| --- | --- | --- | --- | --- |
| Ct range by qPCR | 18S-qPCR | DBP-PCR+ | Microscopic+ | RDT+ |
| <20 | 0 | 0 | 0 | 0 |
| 20 - <25 | 5 | 5 | 0 | 5 |
| 25 - <30 | 17 | 17 | 0 | 7 |
| 30 - <35 | 20 | 18 | 0 | 0 |
| >35 - <38 | 45 | 43 | 0 | 0 |
| Community-based samples (total N= 580) |  |  |  |  |
| <20 | 3 | 3 | 0 | 3 |
| 20 - <25 | 3 | 2 | 0 | 0 |
| 25 - <30 | 1 | 1 | 0 | 0 |
| 30 - <35 | 4 | 4 | 0 | 0 |
| >35 - <38 | 21 | 19 | 0 | 0 |

### Association Between Age, Infection Type, and Symptoms

Age-stratified analysis revealed significant trends in malaria prevalence. Individuals aged 5–15 years had a significantly lower risk of *Pv* infection compared to children under 5 years (OR=0.67, *p*=0.01). It is not surprising that *Pv* was more common in hospital-based participants (10.8%) than those from the community (5.5%), though this difference was not statistically significant (OR=1.16, *p*=0.162). Likewise, mixed infections were also more common in hospital (13.3%) than community samples (7.2%), with a significant two-fold increased risk (OR=2.04, *p*<0.001). Mono-Pv infections were not associated with specific symptoms, while headache was significantly associated with *Pf* infections (OR=3.33, *p*=0.004; Table 3).

**Table 3.** Demographic, Geographic, and Symptom-Based Risk Factors Associated with *Plasmodium vivax*, *Plasmodium falciparum*, and Mixed Infections. This table presents the distribution of malaria infections across various demographic, geographic, and clinical symptom categories. Significant associations (*p*<0.05) were observed for several risk factors. Individuals aged 5–15 years had a significantly lower risk of *P. vivax* infection, while those older than 15 years had a higher risk of *P. falciparum* infection. Hospital-based samples had a higher prevalence of mixed infections, and a lower prevalence of *P. falciparum* compared to community-based cases. Among clinical symptoms, headache was significantly associated with *P. falciparum* infection, whereas no other symptoms showed strong associations. These findings suggest that age, geographic origin, and specific clinical presentations contribute to malaria transmission dynamics and severity.

| Variables | Number of Samples | Total Infections | Odds ratio [95% CI] | *P. vivax* | Odds ratio [95% CI] | *P. falciparum* | Odds ratio [95% CI] | Mixed Infections | Odds ratio [95% CI] |
| --- | --- | --- | --- | --- | --- | --- | --- | --- | --- |
| **Overall** |  |  |  |  |  |  |  |  |  |
|  | 1373 | 50.0% [687] |  | 8.6% [118] |  | 30.7% [421] |  | 10.8% [148] |  |
| **Sex** |  |  |  |  |  |  |  |  |  |
| Female | 59.5% [818] | 58.9% [405] | 1 | 8.9% [73] | 1 | 29.5% [241] | 1 | 11.1% [91] | 1 |
| Male | 40.4% [555] | 41.0% [282] | 0.95 [0.77 to 1.18]<br>**P= 0.636** | 8.1% [45] | 1.11 [0.75 to 1.64]<br>**P= 0.597** | 32.4% [180] | 0.87 [0.69 to 1.09]<br>**P= 0.242** | 10.3% [57] | 1.09 [0.77 to 1.55]<br>**P= 0.617** |
| **Age** |  |  |  |  |  |  |  |  |  |
| <5 | 28.8% [396] | 28.5% [196] | 1 | 5.9% [23] | 1 | 33.2% [128] | 1 | 11.7% [45] | 1 |
| 5-15 | 20.5% [282] | 24.6% [168] | 0.67 [0.49 to 0.91]<br>**P= 0.01\*** | 5.7% [16] | 1.03 [0.53 to 1.98]<br>**P= 0.941** | 43.9% [124] | 0.61 [0.44 to 0.83]<br>**P= 0.002\*\*** | 9.9% [28] | 1.16 [0.71 to 1.91]<br>**P= 0.553** |
| >15 | 50.6% [695] | 46.9% [322] | 1.14 [0.89 to 1.53]<br>**P= 0.314** | 11.2% [78] | 0.49 [0.30 to 0.79]<br>**P= 0.004\*\*** | 24.3% [169] | 1.49 [1.13 to 1.95]<br>**P= 0.004\*\*** | 10.8% [75] | 1.06 [0.72 to 1.57]<br>**P= 0.772** |

| Origin |  |  |  |  |  |  |  |  |  |
| --- | --- | --- | --- | --- | --- | --- | --- | --- | --- |
| Community | 42.2%<br>[580] | 52.2%<br>[303] | 1 | 5.5%<br>[32] | 1 | 39.5%<br>[229] | 1 | 7.2%<br>[42] | 1 |
| Hospital | 57.7%<br>[793] | 48.4%<br>[384] | 1.16 [0.94<br>to 1.44]<br>**P= 0.162** | 10.8%<br>[86] | 0.48<br>[0.32 to<br>0.73]<br>**P=**<br>**0.001\*\*** | 24.2%<br>[192] | 2.04<br>[1.62 to<br>2.58]<br>**P <**<br>**0.0001\*\*** | 13.3%<br>[106] | 0.51 [0.35<br>to 0.74]<br>**P=**<br>**0.0004\*\*** |
| Symptoms |  |  |  |  |  |  |  |  |  |
| Fever | 0.3%<br>[4] | 0.29%<br>[2] | 1 | 25.0%<br>[1] | 1 | 25.0%<br>[1] | 1 | 0 | 1 |
| Chills | 0 | 0 | 1.00 [0.01<br>to 73.27]<br>**P= 1.000** | 0 | 0.43<br>[0.01 to<br>33.59]<br>**P= 0.703** | 0 | 0.43<br>[0.01 to<br>33.59]<br>**P= 0.703** | 0 | 0.11 [0.00<br>to 14.76]<br>**P = 0.378** |
| Abdominal<br>Pain | 0.4%<br>[5] | 0.29%<br>[2] | 1.50 [0.11<br>to 21.31]<br>**P = 0.765** | 0 | 4.71<br>[0.15 to<br>151.49]<br>**P= 0.381** | 0 | 4.71<br>[0.15 to<br>151.49]<br>**P= 0.381** | 40.0%<br>[2] | 0.15<br>[0.01to<br>4.39]<br>**P= 0.275** |
| Headache*<br>(data<br>missing) | 0.8%<br>[11] | 0.73%<br>[5] | 1.20 [0.12<br>to 11.87]<br>**P= 0.876** | 9.1%<br>[1] | 3.33<br>[0.16 to<br>70.91]<br>**P= 0.440** | 9.1%<br>[1] | 3.33<br>[0.16 to<br>70.91]<br>**P= 0.440** | 27.3%<br>[3] | 0.27 [0.01<br>to 6.46]<br>**P= 0.419** |
| Nausea | 0.1%<br>[1] | 0 | 3.00 [0.08<br>to 115.35]<br>**P= 0.555** | 0 | 1.29<br>[0.03 to<br>53.52]<br>**P= 0.895** | 0 | 1.29<br>[0.03 to<br>53.52]<br>**P= 0.895** | 0 | 0.33 [0.00<br>to 25.41]<br>**P= 0.619** |
| Diarrhea | 0.2%<br>[2] | 0 | 5.00 [0.15<br>to 166.59]<br>**P= 0.368** | 0 | 2.14 [0.06<br>to 77.54]<br>**P= 0.677** | 0 | 2.14 [0.06<br>to 77.54]<br>**P= 0.677** | 0 | 0.56 [0.01<br>to 37.56]<br>**P= 0.785** |
| Difficulty<br>Breathing | 0 | 0 | 1.00 [0.01<br>to 73.27]<br>**P= 1.000** | 0 | 0.43<br>[0.01 to<br>33.59]<br>**P= 0.703** | 0 | 0.43<br>[0.01 to<br>33.59]<br>**P= 0.703** | 0 | 0.11 [0.00<br>to 14.76]<br>**P= 0.378** |
| Muscle<br>Pain | 0.4%<br>[5] | 0.44%<br>[3] | 0.67 [0.05<br>to 9.47]<br>**P= 0.765** | 0 | 4.71<br>[0.15 to<br>151.49]<br>**P= 0.381** | 0.20%<br>[1] | 1.33<br>[0.06 to<br>31.12]<br>**P= 0.858** | 40.0%<br>[2] | 0.16 [0.00<br>to 4.39]<br>**P= 0.275** |
| Fatigue | 0.7%<br>[9] | 0.44%<br>[3] | 2.00 [0.18<br>to 22.05]<br>**P= 0.571** | 22.2%<br>[2] | 0.67<br>[0.05 to<br>9.47]<br>**P= 0.765** | 0 | 8.14<br>[0.26 to<br>250.74]<br>**P= 0.230** | 11.1%<br>[1] | 0.63 [0.02<br>to 18.83]<br>**P= 0.789** |

### Genetic polymorphisms of *PvDBP1* and Phylogenic Relatedness

Eight mutations were identified in the *PvDBP1* region II (amino acid 291-460) among 75 *P. vivax* isolates from Cameroon. Two of the mutations occurred at high frequency in the Duffy-negative populations, including I379L observed in 74.1% and E225K in 53.7% of the samples (Figure 4A). Other less frequent mutations included N372K (27.8%), G339D (22.2%), W442C (11.1%), K234E (47.1%), K326E (7.4%), and G417V (5.6%). Variants K326E, G339D, N372K, and I379L were shared with the Thai isolates, though I379L (30%) and G339D (19%) were observed at considerably lower frequencies in Thailand (Figure 4B). Only K326E and N372K were shared between the Cameroonian and Botswana isolates. The Chinese *Pv* were the most polymorphic with 13 variants detected in the binding region, four of which (L333F, N375D, S398T, and T404R) were unique and not observed elsewhere. The remaining nine variants were overlapped with the Sudanese, Ethiopian, and Brazilian isolates. However, none of these variants were detected in the Cameroonian or Botswana *Pv*. The Cameroonian *Pv* shared only N417K with the Brazilian/East African isolates (Figure 4B).

**Figure 4.**
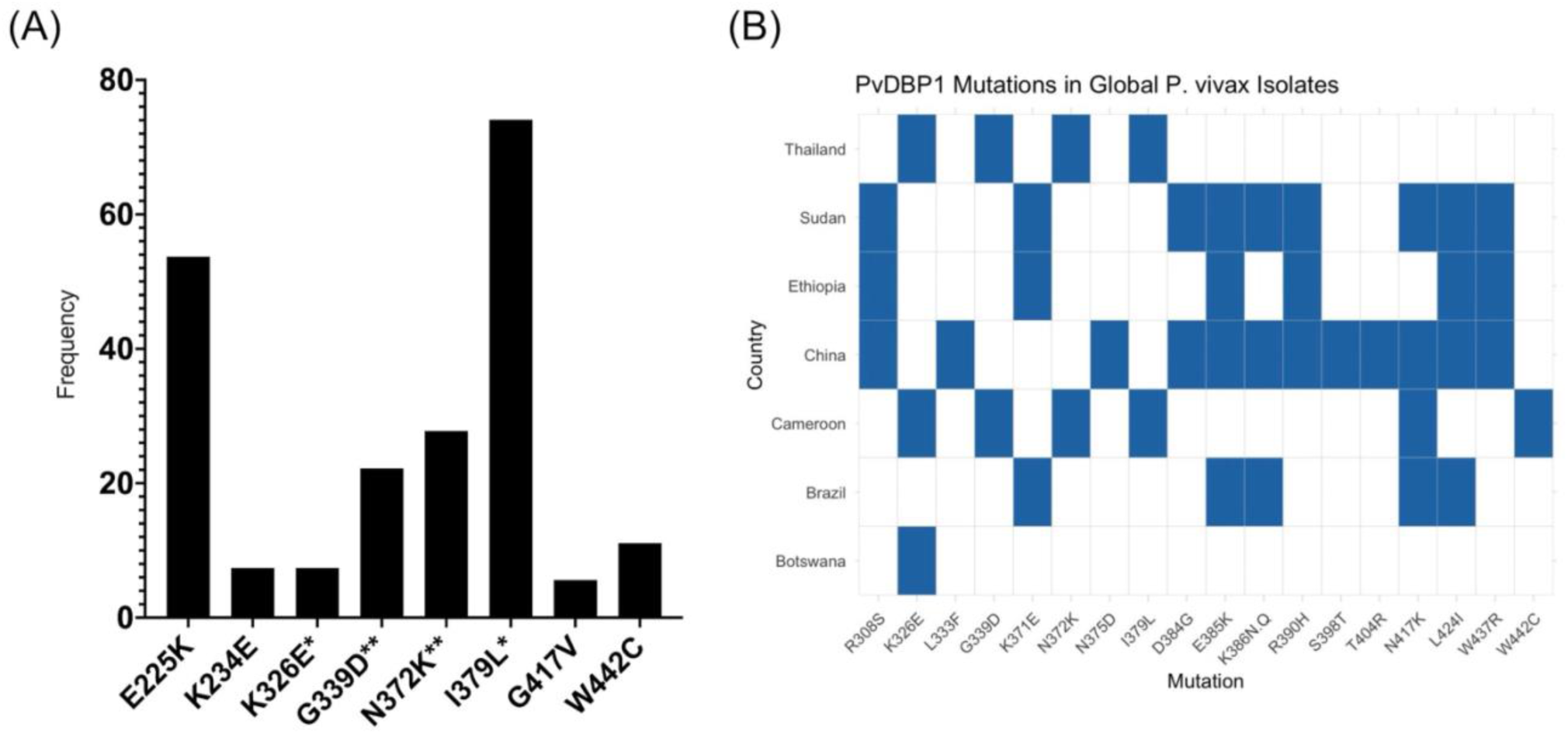
Distribution of DBP Gene Mutations in *P. vivax* Isolates from Cameroon and Global Comparisons: (**A**) The bar chart illustrates the frequency of *Plasmodium vivax* Duffy Binding Protein (DBP) gene mutations in isolates from Cameroon. The mutations E225K, K234E, K326E, G339D, N372K, I379L, G417V, and W442C were observed with varying prevalence, with I379L showing the highest frequency. Mutations marked with an asterisk (*)— K326E, G339D, N372K, and I379L—are also observed in Thailand. (**B**) Comparison of *PvDBP1* Region II mutations observed in *P. vivax* isolates from Cameroon with those reported in geographically and epidemiologically diverse regions, including Thailand and China (Asia), Brazil (South America), Sudan and Ethiopia (East Africa), and Botswana (Southern Africa). Cameroon shared four mutations (K326E, G339D, N372K, and I379L) with Thailand, though the frequencies of I379L and G339D were lower in Thai isolates. K326E and I379L were also found in Botswana. Brazilian and Sudanese/Ethiopian isolates exhibited a broader mutation profile, including several variants not found in Cameroon. Chinese isolates displayed unique mutations such as L333F, S398T, and T404R.

Phylogenetic analysis revealed distinct clustering of *Pv* isolates by geographic region (Figure 5A), with East African and Asian *Pv* showing high genetic relatedness, and South American *Pv* forming a separate clade distinct from the other regions. Within Africa (Figure 5B), most of the isolates from Cameroon (red) form a distinct cluster, sister to those from Botswana (light blue). Two other main clades were observed—one contained most of the Ethiopian (purple) and Ugandan (orange) isolates, and another contained the Ethiopian and Sudanese (green) isolates—indicating the genetic diversity among the African *P. vivax*.

**Figure 5.**
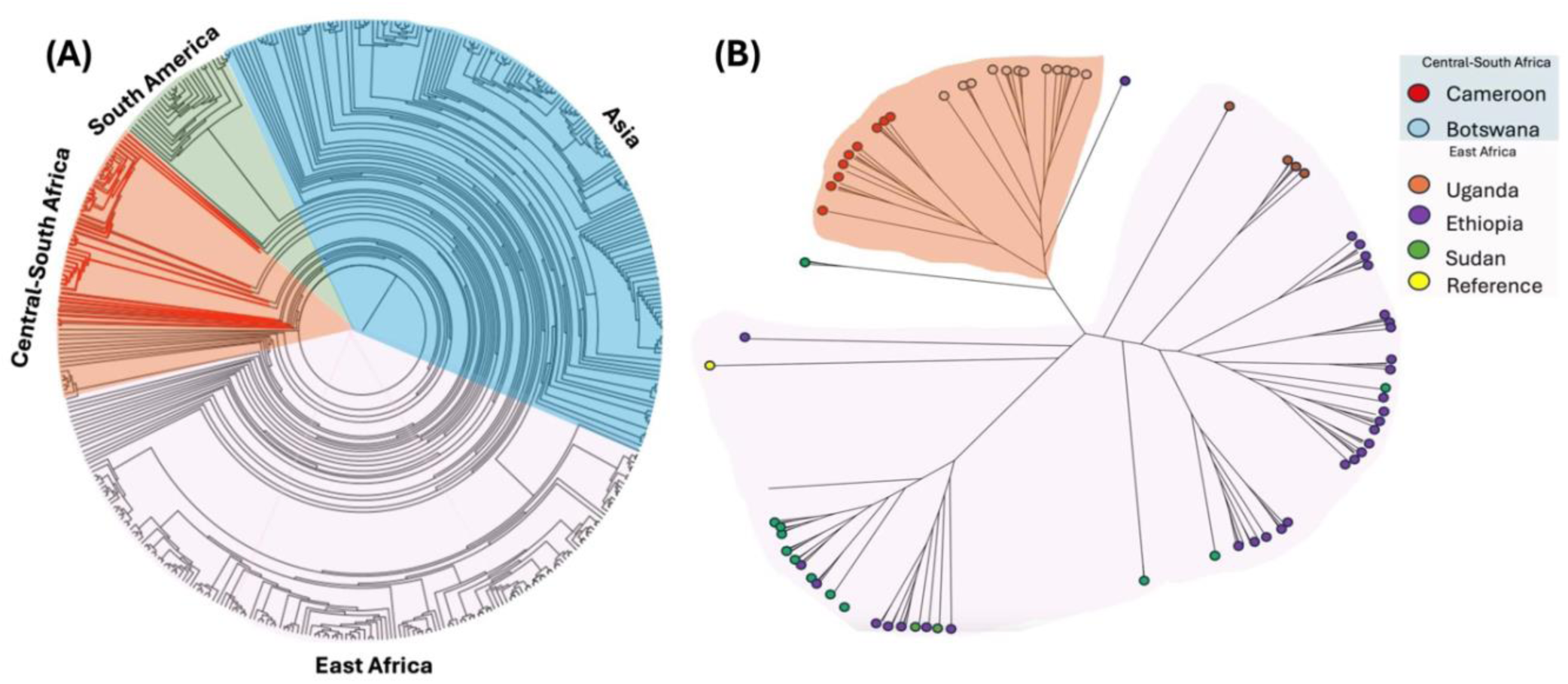
Global and Regional Phylogenetic Relationships of *Plasmodium vivax* Isolates. This figure illustrates the phylogenetic relationships of *Plasmodium vivax* isolates from different geographical regions using whole genome sequence data: (**A**) Displays a circular phylogenetic tree that segregates *P. vivax* populations into major global regions, including Central-South Africa, South America, Asia, and East Africa; (**B**) Presents a radial phylogenetic tree, with color-coded branches corresponding to isolates from Cameroon (red), Botswana (light blue), Uganda (orange), Ethiopia (purple), Sudan (green), and reference genomes (yellow). Both trees demonstrate the genetic diversity and evolutionary divergence of *P. vivax* across different geographical populations, with significant clustering observed among isolates from specific regions. The phylogenetic structure highlights potential gene flow and regional adaptation patterns, particularly in East African populations. These insights are crucial for understanding the evolutionary history and transmission dynamics of *P. vivax* on both global and regional scales.

## Discussion

Molecular assays targeting 18S rRNA and *PvDBP1* revealed a substantial number of *Pv* infections across Cameroon. These findings challenge the longstanding dogma that Duffy-negative individuals are resistant to *Pv*, suggesting instead that the parasite may have well-adapted and circulating widely in Central Africa (7, 16, 18). Geographic and environmental factors strongly influence *Pv* transmission dynamics (19). The southwest region of Cameroon, like Buea at high altitude on Mount Cameroon, provides favorable conditions such as cooler temperatures for the development of *Pv*. Unlike *Pf*, which requires warmer temperatures (18°C and above) mosquito-stage development, *Pv* can survive at 16–18°C, enabling it to transmit in cooler regions (20). High rainfall and flooding in Buea and Bamenda may enhance transmission compared to Bertoua (21). Recent studies suggest that malaria is expanding its endemic range to higher altitudes, including regions such as the Ethiopian highlands (18), Kenya (19), and Colombia (21). In Ethiopia, *P. vivax* has spread into previously malaria-free highland areas with a mono-*P. vivax* infection rate of ∼8%, similar to our findings in this study. It is apparent that *vivax* malaria will increasingly burden the densely populated highlands of Africa and South America. Due to socio-political unrest and large-scale migration in the English-speaking regions of Cameroon since 2016, *Pv* may have readily spread to different parts of Cameroon or beyond. The prevalence of *Pv* infection varied notably by age group. Individuals aged 5–15 years had a significantly lower risk than children under 5 years, suggesting partial immunity or reduced exposure (22). The potential spread and adaptation of *P. vivax* emphasizes the need to consider geographical and environmental heterogeneity when developing malaria control and intervention strategies.

While hospital-based cases had higher parasitemia, some asymptomatic community infections were also shown with high parasitemia, indicating transmission potential. In Ethiopia, *Pv* in Duffy-negative individuals was observed in both febrile patients and asymptomatic carriers (23, 24), and some of these infections were detected with gametocytes (25). These findings emphasize the need for comprehensive surveillance and interventions in both hospital and community settings. The use of microscopy and RDT diagnostics, though convenient, could have vastly misidentified or missed low-density *P. vivax* infections. In our study, some individuals classified as microscopy-negative showed moderate to high parasitemia by qPCR, raising concerns about the diagnostic sensitivity of microscopy and PvLDH-based RDTs, especially in Duffy-negative individuals. Similar microscopy results have been reported in a study from Indonesia(26), where microscopy detected only 5 mixed infections of *P. falciparum* and *P. vivax*, while nested PCR detected 346 cases across various clinical categories, including asymptomatic individuals and those recently treated with antimalarials. Notably, all microscopy results were negative in individuals’ post-treatment, while PCR still detected ongoing parasitemia in most cases(26). Additionally, our findings also suggest that PvLDH-based RDTs may contribute to false-negative results, even in cases of high parasitemia. This could be attributed to defective or degraded test kits, lot variability, or antigenic variation that affects the detection capability of PvLDH in majority Duffy-negative Africa. Molecular tools like qPCR offer superior sensitivity and specificity, particularly in detecting low-density or mixed *P. vivax* infections in Duffy-negative individuals, ensuring both symptomatic and asymptomatic carriers are identified and monitored effectively.

The *PvDBP1* gene encodes a critical ligand that facilitates *P*. *vivax* invasion of human erythrocytes by binding to the Duffy antigen receptor, making it a key molecular marker for both species confirmation and evolutionary analysis. While previous studies in Cameroon have documented the presence of *P. vivax*, this is the first study to utilize *PvDBP1* not only to validate *P. vivax* infections in Duffy-negative individuals, but also to examine genetic variations and phylogenetic relationships of the Cameroonian isolates with global *P. vivax* populations. In the Cameroonian isolates, mutations I379L and E225K occurred at notably high frequencies.

These and several other identified mutations, including K326E, G339D, and N372K overlapped with *P. vivax* populations from Southeast Asia(27). Mutations I379L and G339D occurred at substantially higher frequencies in our dataset than other geographical isolates, suggesting regional adaptation or selective pressures in Central African parasite populations. These findings highlight the need for future functional assays to clarify their potential role in parasite invasion and virulence.

Previous studies indicated striking similarity of the *Pv* genome between Asian and East African parasite populations with high identity-by-descent (20). Our prior research showed no clear genetic distinction between *Pv* in Duffy-negative and Duffy-positive hosts, indicating shared ancestry and gene flow (16, 28). This study further shows that Cameroonian and Botswanan *Pv* form a distinct cluster, separate from East African isolates, likely due to ecological adaptation and vector diversity. For instance, *Pv* parasites show strong compatibility with local mosquito vectors (29). In Cameroon and Benin, *Anopheles coluzzii* was identified as a competent vector for *Pv* (30), suggesting its potential significance in sustaining transmission in West/Central Africa (31). By contrast, in East Africa, *An. arabiensis* and *An. funestus* are primary vectors for malaria transmission. While *An. arabiensis* is more commonly associated with *Pf*, there is evidence suggesting its potential role in transmitting *Pv* (32–34). Such vector differences may drive *Pv* population divergence between regions.

Apart from vector-parasite co-evolution, host genetic factors may also shape parasite population structure. Evidence of severe population bottlenecks followed by rapid expansion in human *Pv*, along with gene pseudogenization (e.g., RBP2d, RBP3) supports host-specific adaptation in *Pv* (35). East Africa, with a larger Duffy-positive population, likely harbors an older, more diverse *Pv* gene pool; whereas Central/Southern Africa may represent a portion of *Pv* parasites recently introduced from East Africa or constrained populations shaped by bottlenecks and the predominance of Duffy-negative hosts (35, 36). The observed genetic relatedness between Cameroonian and Botswanan isolates suggests possible recent gene flow. The Bantu expansion is the most significant and well-studied migration event between Central and Southern Africa, where Bantu-speaking people migrated from West-Central Africa, eventually settling in much of the southern half of the continent and interacting with indigenous groups like the Khoisan, around 3,000 to 5,000 years ago (37, 38). This migration event could have shaped the parasite population structure.

This study has a few limitations. First, samples and related metadata were collected at a single timepoint and thus restricts our ability to assess *Pv* transmission over time. Also, the genetic analysis based on *PvDBP1* alone does not reflect evolutionary changes at the genome level for *Pv* in Duffy-negative individuals. The geographic representation of African *Pv* isolates, particularly samples from West Africa, is limited in the phylogenetic analysis. The apparent genetic distinctiveness of Central African isolates may change with broader sampling across the continent. Our ongoing investigations use a genome-based approach together with spatial analyses to determine source-sink dynamics of *Pv* infections, with the goal to unravel the origin and adaptation of *Pv* in Africa.

To conclude, the detection of several *Pv* cases in asymptomatic individuals underscores its potential for silent transmission, emphasizing the need for enhanced surveillance in both community and clinical settings. PvLDH-based RDTs may contribute to false-negative diagnostic results, even in cases of high parasitemia. Molecular tools like qPCR offer superior sensitivity and specificity, particularly in detecting low-density or mixed *Pv* in Duffy-negative individuals. Our present findings call for tailored malaria control strategies that consider both geographical/environmental heterogeneity and the unique features of *Pv* in Duffy-negative populations.

## Data Availability

All data produced in the present work are contained in the manuscript.

## Acknowledgments

We thank the dedicated field team in Cameroon for their invaluable efforts in sample collection, processing, and logistical support; the communities and hospitals for their support and willingness to participate in this research; and our collaborators and supporting institutions for their contributions and insights, which have significantly enriched this research.

## Funding

This research was supported by NIH/NIAID R21AI190606. The funding body had no role in the study design, data collection, analysis, manuscript preparation, or decision to publish.

## Disclosure of Interest

The authors declare no conflict of interest.

## Notes

### Competing Interest Statement

The authors have declared no competing interest.

### Author Declarations

Ethics committee/IRB of Drexel University gave ethical approval for this work.

### Summary of Updates

This version of the manuscript has been substantially revised to improve clarity scientific rigor and alignment with the journals scope New Visualizations: Figures 2 and 4 were revised for clarity including enhanced legends and standardized color schemes A supplemental figure was added to show the geographic distribution of cases by Duffy phenotype

## References

1. Dieng CC, Gonzalez L, Pestana K, Dhikrullahi SB, Amoah LE, Afrane YA, et al. Contrasting Asymptomatic and Drug Resistance Gene Prevalence of Plasmodium falciparum in Ghana: Implications on Seasonal Malaria Chemoprevention. Genes (Basel). 2019 Jul 16;10(7).

2. King CL, Adams JH, Xianli J, Grimberg BT, McHenry AM, Greenberg LJ, et al. Fy(a)/Fy(b) antigen polymorphism in human erythrocyte Duffy antigen affects susceptibility to Plasmodium vivax malaria. Proc Natl Acad Sci U S A. 2011 Dec 13;108(50):20113–8.

3. Ngum NH, Fakeh NB, Lem AE, Mahamat O. Prevalence of malaria and associated clinical manifestations and myeloperoxidase amongst populations living in different altitudes of Mezam division, North West Region, Cameroon. Malar J. 2023 Jan 19;22(1):20.

4. Gautret P, Legros F, Koulmann P, Rodier M, Jacquemin J-L. Imported Plasmodium vivax malaria in France: geographical origin and report of an atypical case acquired in Central or Western Africa. Acta tropica. 2001;78(2):177–81.

5. Mangoni ED, Severini C, Menegon M, Romi R, Ruggiero G, Majori G. Case report: An unusual late relapse of Plasmodium vivax malaria. American Journal of Tropical Medicine and Hygiene. 2003;68(2):159–60.

6. Russo G, Faggioni G, Paganotti GM, Djeunang Dongho GB, Pomponi A, De Santis R, et al. Molecular evidence of Plasmodium vivax infection in Duffy negative symptomatic individuals from Dschang, West Cameroon. Malar J. 2017 Feb 14;16(1):74.

7. Djeunang Dongho GB, Gunalan K, L’Episcopia M, Paganotti GM, Menegon M, Efeutmecheh Sangong R, et al. Plasmodium vivax Infections Detected in a Large Number of Febrile Duffy-Negative Africans in Dschang, Cameroon. Am J Trop Med Hyg. 2021 Jan 12;104(3):987–92.

8. Ntumngia FB, McHenry AM, Barnwell JW, Cole-Tobian J, King CL, Adams JH. Genetic variation among Plasmodium vivax isolates adapted to non-human primates and the implication for vaccine development. Am J Trop Med Hyg. 2009 Feb;80(2):218–27.

9. Seraphin EH, Lagarde BJ, Richard P, Jules N, Farrick NO. Diversity, structure and health of a cocoa based agroforest system in the Humid dense forest, East Cameroon. International Journal of Biodiversity andConservation. 2021:165.

10. Akenji T, Ntonifor N, Kimbi H, Abongwa E, Ching J, Anong D, et al. The epidemiology of malaria in Bolifamba, a rural setting of Mount Cameroon: Seasonal variations in parasitological indices of transmission [MIM-TA-141660]. ACTA TROPICA; 2005: ELSEVIER SCIENCE BV PO BOX 211, 1000 AE AMSTERDAM, NETHERLANDS; 2005. p. S295–S6.

11. Fru-Cho J, Bumah VV, Safeukui I, Nkuo-Akenji T, Titanji VP, Haldar K. Molecular typing reveals substantial Plasmodium vivax infection in asymptomatic adults in a rural area of Cameroon. Malar J. 2014 May 3;13:170.

12. Miguel RB, Coura JR, Samudio F, Suárez-Mutis MC. Evaluation of three different DNA extraction methods from blood samples collected in dried filter paper in Plasmodium subpatent infections from the Amazon region in Brazil. Rev Inst Med Trop Sao Paulo. 2013;55(3).

13. Lo E, Yewhalaw D, Zhong D, Zemene E, Degefa T, Tushune K, et al. Molecular epidemiology of Plasmodium vivax and Plasmodium falciparum malaria among Duffy-positive and Duffy-negative populations in Ethiopia. Malar J. 2015 Feb 19;14:84.

14. Abagero BR, Rama R, Obeid A, Tolossa T, Legese F, Lo E, et al. Detection of Duffy Blood Group Genotypes and Submicroscopic Plasmodium Infections Using Molecular Diagnostic Assays in Febrile Malaria Patients. Res Sq. 2023 Dec 6.

15. Gunalan K, Lo E, Hostetler JB, Yewhalaw D, Mu J, Neafsey DE, et al. Role of Plasmodium vivax Duffy-binding protein 1 in invasion of Duffy-null Africans. Proc Natl Acad Sci U S A. 2016 May 31;113(22):6271–6.

16. Lo E, Russo G, Pestana K, Kepple D, Abagero BR, Dongho GBD, et al. Contrasting epidemiology and genetic variation of Plasmodium vivax infecting Duffy-negative individuals across Africa. International Journal of Infectious Diseases. 2021;108:63–71.

17. Ménard D, Barnadas C, Bouchier C, Henry-Halldin C, Gray LR, Ratsimbasoa A, et al. Plasmodium vivax clinical malaria is commonly observed in Duffy-negative Malagasy people. Proc Natl Acad Sci U S A. 2010 Mar 30;107(13):5967–71.

18. Ketema T, Bacha K, Getahun K, Portillo HAD, Bassat Q. Plasmodium vivax epidemiology in Ethiopia 2000-2020: A systematic review and meta-analysis. PLoS Negl Trop Dis. 2021 Sep;15(9):e0009781.

19. Shanks GD, Hay SI, Omumbo JA, Snow RW. Malaria in Kenya’s western highlands. Emerg Infect Dis. 2005 Sep;11(9):1425–32.

20. Benavente ED, Manko E, Phelan J, Campos M, Nolder D, Fernandez D, et al. Distinctive genetic structure and selection patterns in Plasmodium vivax from South Asia and East Africa. Nat Commun. 2021 May 26;12(1):3160.

21. Siraj AS, Santos-Vega M, Bouma MJ, Yadeta D, Ruiz Carrascal D, Pascual M. Altitudinal changes in malaria incidence in highlands of Ethiopia and Colombia. Science. 2014 Mar 7;343(6175):1154-8.

22. Wamae K, Wambua J, Nyangweso G, Mwambingu G, Osier F, Ndung’u F, et al. Transmission and Age Impact the Risk of Developing Febrile Malaria in Children with Asymptomatic Plasmodium falciparum Parasitemia. J Infect Dis. 2019 Feb 23;219(6):936–44.

23. Bradley L, Yewhalaw D, Hemming-Schroeder E, Jeang B, Lee MC, Zemene E, et al. Comparison of Plasmodium Vivax Infections in Duffy Negatives From Community and Health Center Collections in Ethiopia. Res Sq. 2023 Oct 3.

24. Bradley L, Yewhalaw D, Hemming-Schroeder E, Jeang B, Lee MC, Zemene E, et al. Epidemiology of Plasmodium vivax in Duffy negatives and Duffy positives from community and health centre collections in Ethiopia. Malar J. 2024 Mar 14;23(1):76.

25. Singh K, Mukherjee P, Shakri AR, Singh A, Pandey G, Bakshi M, et al. Malaria vaccine candidate based on Duffy-binding protein elicits strain transcending functional antibodies in a Phase I trial. NPJ Vaccines. 2018;3:48.

26. Deo DA, Herningtyas EH, Intansari US, Perdana TM, Murhandarwati EH, Soesatyo M. Difference between Microscopic and PCR Examination Result for Malaria Diagnosis and Treatment Evaluation in Sumba Barat Daya, Indonesia. Trop Med Infect Dis. 2022 Jul 29;7(8).

27. Tapaopong P, da Silva G, Chainarin S, Suansomjit C, Manopwisedjaroen K, Cui L, et al. Genetic diversity and molecular evolution of Plasmodium vivax Duffy Binding Protein and Merozoite Surface Protein-1 in northwestern Thailand. Infect Genet Evol. 2023 Sep;113:105467.

28. Kepple D, Hubbard A, Ali MM, Abargero BR, Lopez K, Pestana K, et al. Plasmodium vivax From Duffy-Negative and Duffy-Positive Individuals Share Similar Gene Pools in East Africa. J Infect Dis. 2021 Oct 28;224(8):1422–31.

29. Joy DA, Gonzalez-Ceron L, Carlton JM, Gueye A, Fay M, McCutchan TF, et al. Local adaptation and vector-mediated population structure in Plasmodium vivax malaria. Mol Biol Evol. 2008 Jun;25(6):1245–52.

30. Feufack-Donfack LB, Sarah-Matio EM, Abate LM, Bouopda Tuedom AG, Ngano Bayibéki A, Maffo Ngou C, et al. Epidemiological and entomological studies of malaria transmission in Tibati, Adamawa region of Cameroon 6 years following the introduction of long-lasting insecticide nets. Parasit Vectors. 2021 May 8;14(1):247.

31. Ossè RA, Tokponnon F, Padonou GG, Glitho ME, Sidick A, Fassinou A, et al. Evidence of Transmission of Plasmodium vivax 210 and Plasmodium vivax 247 by Anopheles gambiae and An. coluzzii, Major Malaria Vectors in Benin/West Africa. Insects. 2023 Feb 25;14(3).

32. Massebo F, Balkew M, Gebre-Michael T, Lindtjørn B. Entomologic inoculation rates of Anopheles arabiensis in southwestern Ethiopia. Am J Trop Med Hyg. 2013 Sep;89(3):466–73.

33. Eligo N, Wegayehu T, Pareyn M, Tamiru G, Lindtjørn B, Massebo F. Anopheles arabiensis continues to be the primary vector of Plasmodium falciparum after decades of malaria control in southwestern Ethiopia. Malar J. 2024 Jan 9;23(1):14.

34. Ogola EO, Odero JO, Mwangangi JM, Masiga DK, Tchouassi DP. Population genetics of Anopheles funestus, the African malaria vector, Kenya. Parasit Vectors. 2019 Jan 8;12(1):15.

35. Loy DE, Plenderleith LJ, Sundararaman SA, Liu W, Gruszczyk J, Chen YJ, et al. Evolutionary history of human Plasmodium vivax revealed by genome-wide analyses of related ape parasites. Proc Natl Acad Sci U S A. 2018 Sep 4;115(36):E8450–e9.

36. Abebe A, Dieng CC, Dugassa S, Abera D, Shenkutie TT, Assefa A, et al. Genetic differentiation of Plasmodium vivax duffy binding protein in Ethiopia and comparison with other geographical isolates. Malar J. 2024 Feb 23;23(1):55.

37. Campbell MC, Tishkoff SA. The evolution of human genetic and phenotypic variation in Africa. Curr Biol. 2010 Feb 23;20(4):R166–73.

38. Campbell MC, Hirbo JB, Townsend JP, Tishkoff SA. The peopling of the African continent and the diaspora into the new world. Curr Opin Genet Dev. 2014 Dec;29:120–32.

