## Supporting Information for "Epidemiological Insights and Duffy Binding Protein Evolution of *Plasmodium vivax* in Duffy-Negative Cameroonians"

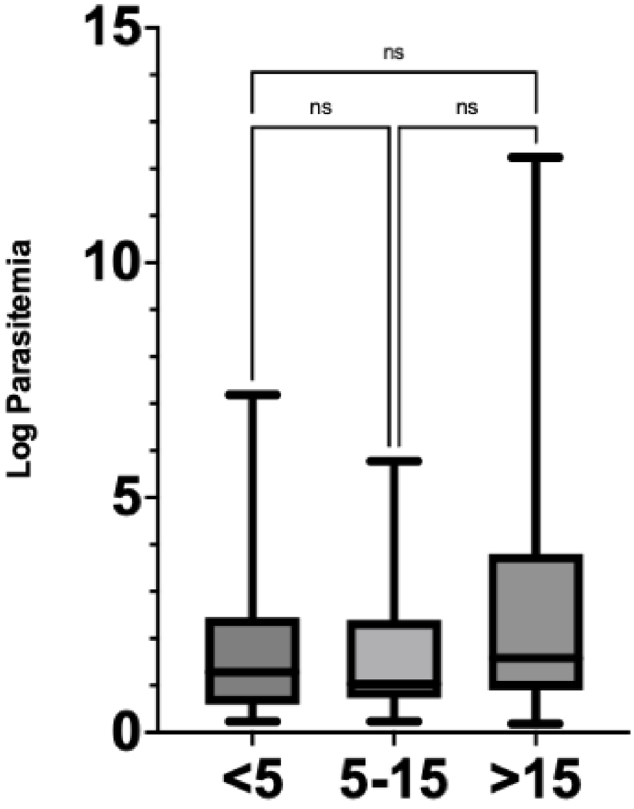

**S1 Fig.** This figure compares parasitemia levels across different age groups (<5, 5-15, >15 years) with no significant differences observed between the age groups (ns).

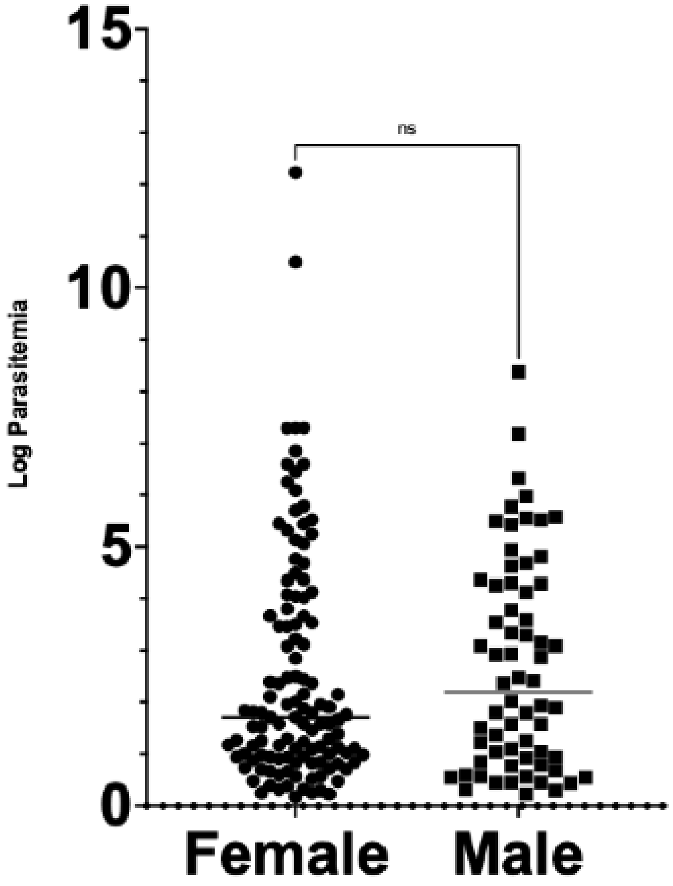

**S2 Fig.** This figure examines the parasitemia levels between female and male participants, with no significant differences observed (ns).

| *Plasmodium Falciparum* **Primers** | |
|---|---|
| Forward | TAT TGC TTT TGA GAG GTT TTG TTA CTT TG |
| Reverse | ACC TCT GAC ATC TGA ATA CGA ATG C |
| ***Plasmodium vivax* Primers** | |
| Forward | GCT TTG TAA TTG GAA TGA TGG GAA T |
| Reverse | ATG CGC ACA AAG TCG ATA CGA AG |

**S1 Table.** 18S SYBR Primers
